# Five Year Pediatric Use of a Digital Wearable Fitness Device: Lessons from a Pilot Case Study

**DOI:** 10.1101/2020.10.21.20215491

**Authors:** Kimayani D. Butte, Amir Bahmani, Atul J. Butte, Xiao Li, Michael P. Snyder

## Abstract

**Objectives:** Wearable fitness devices are increasingly used by the general population, with new applications being proposed and designed for healthy adults as well as adults with chronic diseases. Fewer, if any, studies of these devices have been conducted in healthy adolescents and teenagers, especially over a long period of time. The goal of this work was to document the successes and challenges involved in 5 years of a wearable fitness device use in a pediatric case study.

**Materials and Methods:** Comparison of five years of step counts and minutes asleep from a teenaged girl and her father.

**Results:** At 60 months, this may be the longest reported pediatric study involving a wearable fitness device, and the first simultaneously involving a parent and a child. We find step counts to be significantly higher for both the adult and teen on school/work days, along with less sleep. The teen walked significantly less towards the end of the 5 year study. Surprisingly, many of the adult’s and teen’s sleeping and step counts were correlated, possibly due to coordinated behaviors.

**Discussion:** We end with several recommendations for pediatricians and device manufacturers, including the need for constant adjustments of stride length and calorie counts as teens are growing.

**Conclusion:** With periodic adjustments for growth, this pilot study shows these devices can be used for more accurate and consistent measurements in adolescents and teenagers over longer periods of time, to potentially promote healthy behaviors.

## Background and Significance

Wearable fitness devices are increasingly used by the general population. A recent study estimates that 19% of Americans currently use a wearable fitness device ^1^ and an additional 15% of Americans stated they no longer used a fitness tracker, which raises a question on their long-term use. Forty million new wearable devices were sold in 2017 ^2^, and the company Fitbit sold approximately 13.5 million new Fitbit devices in 2019 ^3^.

While many consumers likely use these devices to track general health and wellness parameters, there have been several attempts to discover medical utility for these devices. Gresham, et al, recently showed in a study of 37 patients with cancer that wearable fitness devices could accurately capture stair climbing and steps with enough accuracy to correlate with traditional performance status measures assessed by health providers ^4^. Similarly, Speier, et. al., showed that wearable fitness devices could be very useful in monitoring a patient’s health remotely in order to diagnose health problems more quickly ^5^. The study was conducted for only 90 days with 186 participants with ischemic heart disease. Li, et. al., demonstrated that fitness trackers can detect illnesses caused by infectious diseases such as Lyme and respiratory viral infections as well as other health conditions ^6^. Finally, Rose, et. al., have used fitness trackers to detect heart conditions, such as atrial fibrillation ^7^.

One of the longest studies on the use of wearable fitness devices was by Jakicic, et al, who studied their utility for weight loss in young adults (ages 18 to 35 years) ^8^. The study randomized 471 participants, of which 351 were still with the study and provided their updated weights at 24 months. The study showed a significant decrease in weight over time, but surprisingly with less weight loss in the intervention group using the wearable fitness devices. Also interestingly, nearly a quarter of the participants did not complete the study over 2 years. Despite these setbacks, the research use of wearable fitness devices is forecast to continue to expand, with over 500 biomedical publications specifically mentioning Fitbit. With the flagship NIH *All of Us* Research Program studies now adopting wearable fitness data along with electronic health record data and genomic data, research using these devices is likely to increase ^9^.

To date, the vast majority of research on wearable fitness devices has been conducted in adults. Fewer studies have been performed in the pediatric population, though companies are now targeting sales growth of digital wearable devices in this population ^10^. The few pediatric studies that have been performed have targeted cohorts with specific diseases. Bian et al looked at self-reported sleep quality from 22 participants with asthma as well as self-reported symptoms of asthma, and compared the reports to the participants’ Fitbit data to show that Fitbit sleep quality is lower when more asthma symptoms occur ^11^. Voss, et al., conducted a study with 40 participants ages 10-18 with congenital heart disease. The study assessed the validity of the Fitbit step count by testing the wrist-worn Fitbit Charge HR against the hip-worn ActiGraph accelerometer and found that the Fitbits recorded more steps than the accelerometer^12^. The study also found that daily Fitbit step counts of over 12,500 steps would meet commonly promoted physical activity guidelines of over 60 minutes of activity per day. A similar study by Miropolsky, et al., on 13 young adult cancer survivors between ages 20 and 39 years suggested a Fitbit device could provide major motivation to engage in physical activity ^13^.

Studies on healthy adolescents and teenagers have been even more rare. Kerner, et al. studied one-hundred participants from two schools using Fitbit devices for 8 weeks, along with the Fitbit app. They found using Fitbits can initially encourage adolescents (ages 13-14) to exercise, but the participants were eventually discouraged potentially because Fitbit might be setting unrealistic goals ^14^. The non-personalized goal of 10,000 steps per day made participants feel unmotivated and lazy if they did not achieve that goal, which discouraged them from exercising^14^. Also, the competitive aspect of the Fitbit app, such as the leaderboard rankings for who got the most steps, discouraged those who did not get many steps and sometimes demotivated the participants who participated in exercise just to get more steps than they usually do. Finally, the authors did not report on the actual success rate of how many students used the Fitbit for the full 8 weeks.

These studies show that using a wearable fitness device could be useful for tracking some health indicators from pediatric-aged individuals, without needing regular physician evaluation. Almost all of these studies involved the participants using devices for 8 weeks or fewer, a period too short for many health indicators to significantly improve. Health benefits from wearable fitness devices might be expected to require longer-term use which could lead to new discoveries in long-term health effects of exercise on medical conditions and general health.

## Objectives

The goal of this work was to document the successes and challenges involved in 5 years of a wearable fitness device use in a pediatric case study. The findings from this study may have implications for both encouraging healthy behavior in adolescents as well as recommendations for wearable device manufactures to improve adoption by adolescents and adults.

## Materials and Methods

Ethics: Two authors (including the lead author) collected their own data as citizen-scientists, with their own devices that they obtained commercially through Amazon.com, initiated the analyses, then approached the Stanford investigators to enhance the analyses, and both of these authors contributed to writing this manuscript. This citizen-science research project was initiated outside of an academic institution, was run by the two authors with their own data, had no Federal funding, and the research is not related to an FDA or EPA-regulated product, so as such does not need to undergo Institutional Review Board review, as per Resnik ^15^. Regardless, the two data contributors joined Stanford University research protocol 56378 approved by the Stanford University Institutional Review Board, specifically allowing participants at or over age 13 years to share their past and current Fitbit measurement data with Stanford investigators for research purposes, with informed consent.

Two Fitbit One devices were purchased on January 1, 2013 and activated shortly thereafter. The Fitbit One was designed to track steps walked along with pace, stairs climbed, sleep duration and activity. Two participants (and co-authors here) simultaneously started to use the devices to track these measurements. The female participant started use at age 10 years and 4 months, and continued through her teenage years. The adult male participant (father of the younger individual) started use at age 43 years and 9 months. Neither had any significant prior medical history at the initiation of use. Both intended the use of these devices for improving and maintaining general health and wellness. The adult also intended to use the device to increase his walking and help in weight loss. While these devices (or their subsequent versions) have still been in use since January 1, 2013, this analysis only covers the 60 months of use after a pattern of regular consistent use was seen, starting on June 1, 2013. In June 2018, both participants elected to study their data together for research.

Fitbit enables the downloading of raw level device data, by registering through their Application Programming Interface (API). Using the API ID number that Fitbit provided, a short program was written in R to access the Fitbit data, serially downloading blocks of daily step and sleep data representing every 100 days, to cover the entire 60 months. No outlier measurements were removed. Sleep amounts in minutes are assigned to the wake up day. Weekend nights were defined as those leading into a Saturday or Sunday, which are days with no school. School days were defined as weekdays that were not within a set of five long holiday breaks: one week mid-winter break in February, one week spring break in March or April, ten week summer break in June-August, Thanksgiving one week break in November, and two week winter break between December and January.

Height data for the teen, needed to calculate stride length, was downloaded from her own electronic medical records, with height measurements made and documented by a physician assistant or pediatrician.

Analysis was performed using Google Sheets, Minitab Express, R [version 4.0.2 (2020-06-22)], and RStudio [version 1.3.1073]. R packages used include ggplot2 [version 3.3.2], tidyverse [version 1.3.0], and corrr [version 0.4.2].

## Results

We wanted to examine the patterns and challenges associated with long term adolescent use of wearable fitness data. Sixty months of step and sleep data, collected from the Fitbit wearable fitness devices of a father and daughter (Table 1) were downloaded and analyzed. School for the teen and work for the adult was generally in session from Monday morning through Friday afternoon and the teen’s school year consistently ran from late August to early June. For the first year of the study, the teen had to was required to complete a “mile run” once per year. In the subsequent 3 years in middle school, the teenager was required to complete a “mile run” once every month. In the final year of the study (which was during high school), there were no daily physical education classes, only lower intensity yoga without mandatory walking or running.

**Table 1:** Data collected over 60 months from the teen and adult.

|  | **Teen Female** | **Adult Male** |
| --- | --- | --- |
| Starting age | 10 years 4 months (ending 4th grade) | 43 years 9 months |
| Available step measurements (days) | 1566 (85.8%) | 1823 (99.8%) |
| Missing step measurements (days) | 260 (14.2%) | 3 (0.16%) |
| Available sleep measurements (days) | 828 (45.3%) | 1548 (84.8%) |
| Missing sleep measurements (days) | 998 (54.7%) | 278 (25.2%) |
| Steps per day, mean (standard deviation) | 6568.3 (3685.9) | 7757.2 (2850.7) |
| Minutes sleep per night, mean (standard deviation) | 482.6 (112.1) | 362.5 (85.7) |

The teen was noted to have more missing measurements than the adult, including a 193 day gap in measurements in 2016. However, more than 85% of the possible 1826 days of step counting were available for both individuals. Fewer sleep measurements were made than step measurements by both individuals, likely due to the need to remember to manually activate and deactivate the Fitbit One sleep timer before and after sleeping.

With over 1500 days of step counts available for both individuals, some clear differences are notable. For the teen, step counts significantly dropped over the five years (Figure 1A, negative correlation of date versus steps r = −0.517, p = 1.20 x 10^−107^), whereas step counts only slightly dropped the same period for the adult (Figure 1B, negative correlation of date versus steps r = - 0.066, p = 0.005). On average, the adult walked significantly more steps than the teen over the 5 years (adult mean 7757.2, standard deviation 2850.7; teen mean 6568.3, standard deviation 3685.9, t-test p < 2.2 x 10^−16^). Interestingly, both individuals walked significantly less on weekend days (Sundays and Saturdays) than on weekdays, but the difference was more pronounced in the teen (Figure 1C and 1D, teen t-test p < 2.2 x 10^−16^, adult t-test p = 0.006).

**Figure 1:**
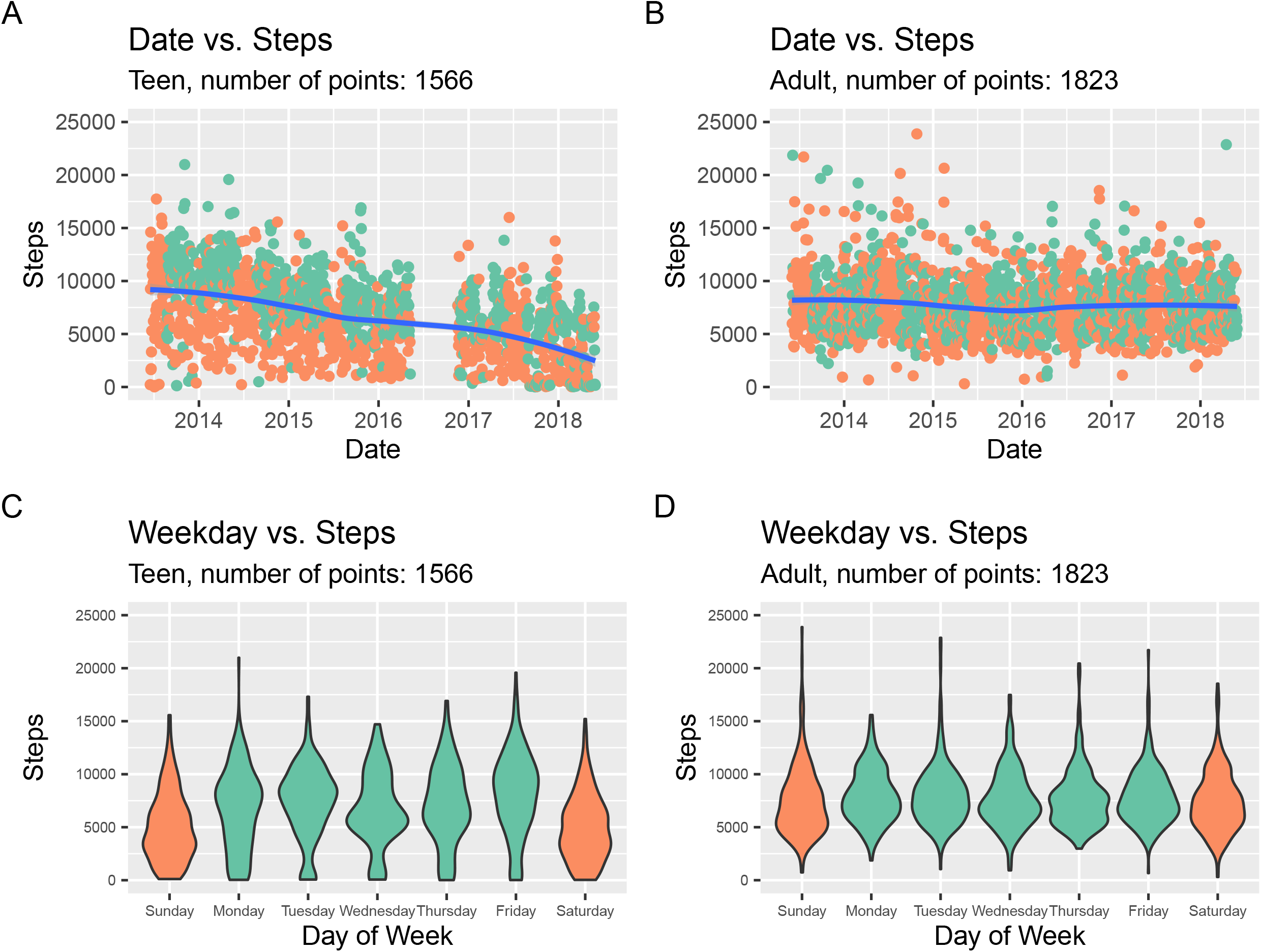
A. and B. Steps per day over time for the teenager and adult, respectively, with green representing school days and orange as non-school days (i.e. weekends and weeklong holidays). C. and D. Steps per day on each day of the week for the teenager and adult, respectively, with green representing weekdays and orange as weekends (Saturday and Sunday).

We further investigated the teen’s step counts. Using six years of school calendars, we determined the specific dates of five yearly recurring holidays (one week mid-winter break in February, one week spring break in March or April, ten week summer break from June through August, Thanksgiving one week break in November, and two week winter break from between December and January). Approximately 30% of the available step count measurements could be classified as occurring during one of these holidays (teen: 472 of 1566 measurements; adult: 571 of 1823 measurements). The teen walked significantly more on school days compared to non-school days (combining holiday and weekend days, Figure 2A, 2105.9 more steps on average, t-test p = 8.17 x 10^−31^). Interestingly, the adult showed no significant difference in step counts between school days and non-school days. The teen made fewest steps in February and March compared to August through October (Figure 2B), and there was significant variability in the steps across the months (Chi-square test p = 3.71 x 10^−67^). To test the effect of seasonality, we fit a linear regression model on the teen’s step counts, with parameters representing whether a day was a weekend, in one of the five holiday periods, the year of the study (first through fifth), and the month of the year. All of these parameters were highly significant in the fit regression model (Table 2).

**Table 2:** Linear regression model fit on the 1826 step counts from the teen, with variables representing whether a day was a weekend, in one of the five holiday periods, the year of the study (first through fifth), and the month of the year.

| Variable in linear regression model | Coefficient | ANOVA p-value |
| --- | --- | --- |
| Intercept | 11215.0 |  |
| Day is a weekend day | – 2194.91 | $< 2.2 \times 10^{-16}$ |
| Day during one of the five holiday periods | – 996.75 | $< 2.2 \times 10^{-16}$ |
| Every year of the study past the first year<br>(First year 0, second year 1, ...) | – 1299.89 | $1.953 \times 10^{-10}$ |
| Month (1 for January, 12 for December) | – 107.01 | $1.425 \times 10^{-6}$ |

**Figure 2:**
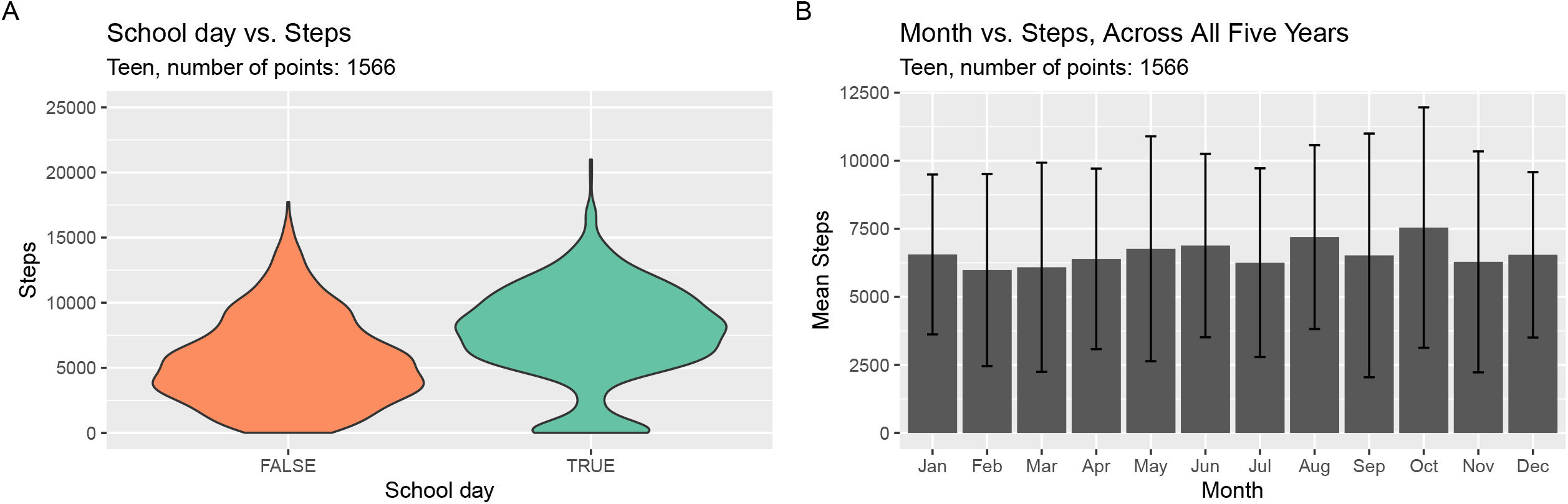
A. Distribution of steps per day for the teenager on school days (green) and non-school days (orange; weekends and weeklong holidays). B. Average and standard deviation of teenager steps for each month of the year.

On average, the teen walked 9103.1 steps per day during the first year of the study, and only 3646.8 steps per day during the final year of the study, or a drop of 60%. The increasing heights for the growing teen were used to better understand the context for this marked drop in step counts. Fourteen height measurements were downloaded for the teen, with measurements available between ages 2 and 16 years. A cubic spline was then fit to these measurements, and a height of 144.9 cm was estimated for the teen on June 1, 2013, the starting date for this study. A height of 162.6 cm (measured close to the date) was used for May 31, 2018, the ending date for this analysis. The teen’s height is estimated to have increased 17.7 cm (or 12.2%) during the analysis period of this study, and thus only a 12.2% increase in stride length over the course of the study ^16,17^.

Fewer days of sleep measurements were available for both individuals, but comparisons were still possible with over 800 nights of data available. The teen slept slightly less towards the end of the 5 years, compared to the start (Figure 3A, correlation of date versus minutes asleep r = - 0.10, p = 0.003). The adult showed no significant change in sleep over the 5 years (Figure 3B, correlation of date versus minutes asleep r = 0.036, p = 0.152, not significant). Similar to the step counts, differences were observed on weekends. Both individuals slept significantly longer over nights when the morning fell on a weekend day. The teen slept an average of 92.7 minutes longer in weekends (Figure 3C, green weekday mean 457.3, orange weekend mean 550.0, t-test p = 7.23 x 10^−29^) whereas the adult slept 61 minutes longer (Figure 3D, green weekday mean 345.5, orange weekend mean 406.3, t-test p = 1.5 x 10^−36^). Overall, the teen slept an average of 120.1 minutes longer per day than the adult.

**Figure 3:**
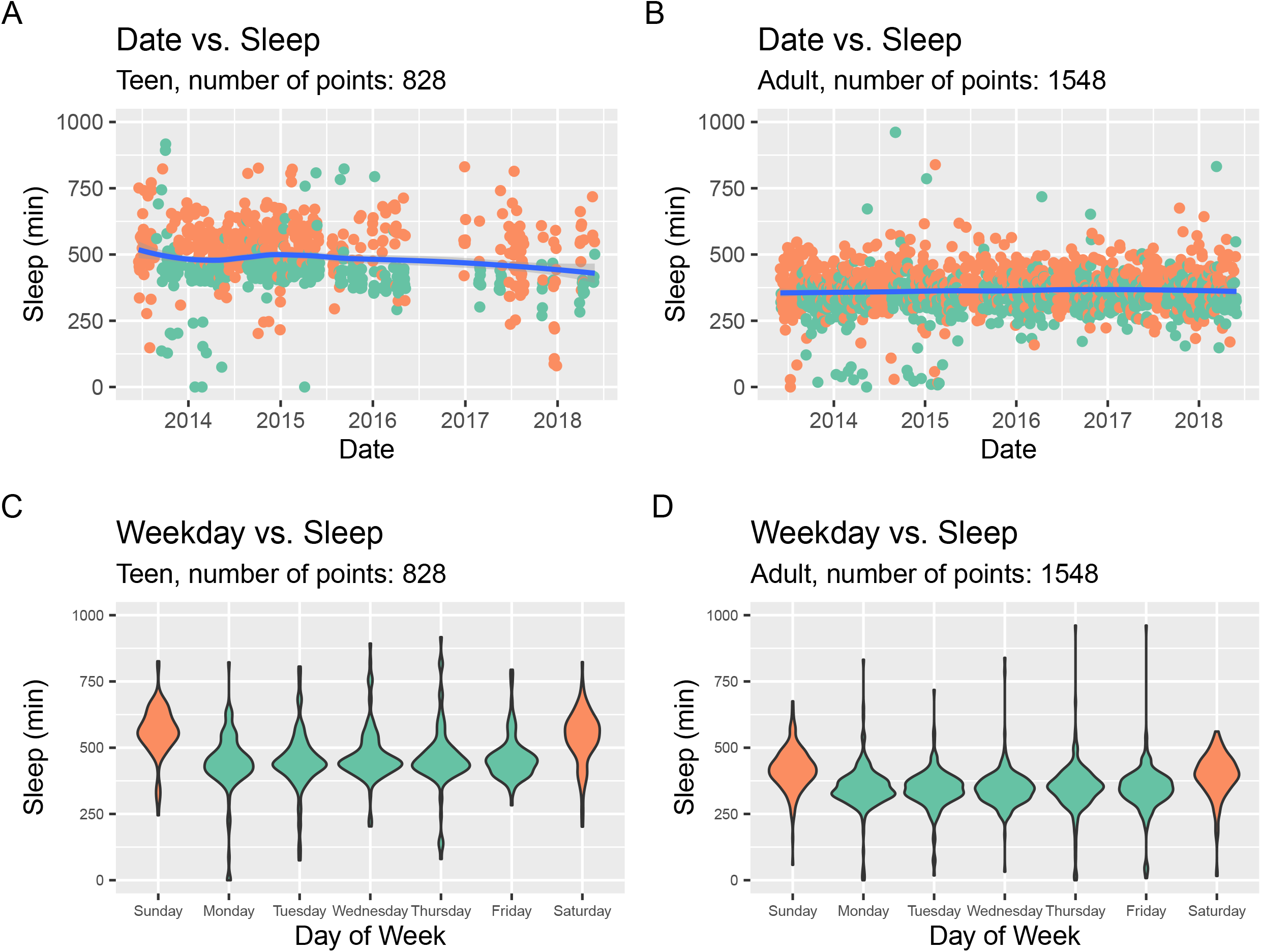
A. and B. Minutes asleep per day over time for the teenager and adult, respectively, with green representing school days and orange as non-school days (i.e. weekends and weeklong holidays). C. and D. Minutes asleep per day on each day of the week for the teenager and adult, respectively, with green representing weekdays and orange as weekends (Saturday and Sunday).

Given the father and daughter living as a family in the same home, we then compared step counts and sleeping on the same days. Surprisingly, the number of steps walked by the teen and adult were correlated (Figure 4A, n = 1589 points, Pearson r = 0.2, p = 4.92 x 10^−16^). The step count correlation was even stronger when considering only the weekend days (n = 449 points, Pearson r = 0.4, p = 1.25 x 10^−18^), suggesting coordinated walking behaviors on the weekend. Interestingly, the amount of sleep was also correlated between the father and daughter (Figure 4B, n = 707 points, Pearson r = 0.23, p = 1.32 x 10^−9^), but much weaker when considering only the weekend days (n = 189 points, Pearson r = 0.17, p = 0.023), suggesting the daughter’s and father’s sleep were less coordinated on the weekends. In general, both the teen and adult slept more and walked less on non-school days (Figure 4A and B, in orange) than on school days (green).

**Figure 4:**
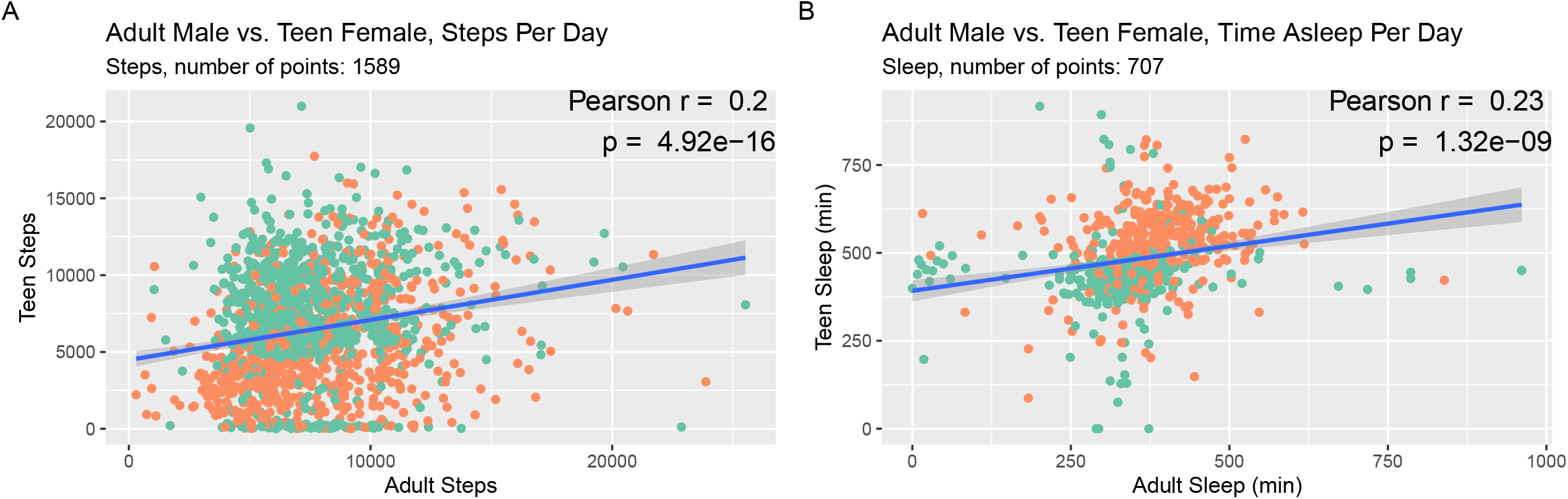
A. Steps per day for the teenager against the number of steps on identical days for the adult, with green representing school days and orange as non-school days. B. Minutes asleep per night for the teenager against the minutes of sleep per night for the adult, on identical days, with green representing school days and orange as non-school days.

## Discussion

We describe here a 5-year longitudinal case study of a wearable fitness device in a teenage female, comparing these measurements to her adult father, using the same type of device during the same time period. To our knowledge, this is the longest reported pediatric study involving a wearable fitness device, and the first simultaneously involving a parent and a child.

We describe five specific findings here. First, the teen appears to be generally walking less than the adult (who was purposefully trying to walk more for weight loss), and progressively walking less during the 5 years. Second, both the adult and teen walk significantly less on weekends and holidays. Third, the teen female sleeps more than the adult and has more variation in sleep, but both the teen and adult sleep more on weekends. Fourth, on average, the teenager slept more than the adult. Sleep differences may have been due to travel by the adult and/or the fact that short naps are often not recorded by these devices, which needed manual triggers to start the timer. Fifth, interestingly, the number of steps walked and minutes slept by the teen and adult are correlated. The correlation in step counts may be due to common activities (e.g. shopping) or longer walks (e.g. hikes) performed together as a family. Similarly, sleep correlations were likely due to shared behavior: if the teen worked late to complete homework, one of the two parents often stayed awake with her.

While we have shown that device use for more than 5 years is possible, we note special efforts that could be taken to enhance continued accurate use in the pediatric setting. First, the number of steps needed to traverse a distance obviously depends on the stride length. If an exact stride length is not entered, Fitbit estimates the length given the sex and height of the user ^18^ using a proprietary formula, which is multiplied by the step count to estimate the distance walked. However, teenagers gain significant height as they go through puberty, and if the stride length is not periodically readjusted, a shorter total distance walked may be misestimated. For example, an average 10-year-old female at the 50%ile for height at 138 cm would have an estimated stride length of 57 cm, while the same female 5 years later at the 50%ile for height at 162 cm would have an estimated stride length of 67 cm or 17.5% longer ^17,19,20^. Thus, 5000 steps walked at age 15 years cover 2.08 miles, whereas 5000 steps for a 10-year-old would cover 1.77 miles. However, in this study, we show that the 60% decrease in step counts seen over the five years greatly exceeds the teen’s 12.2% estimated increase in stride length.

Given that teenager heights change over time, if the teen does not know (or remember) to periodically change their height in the settings, then the Fitbit may miscalculate the number of steps and thus distance walked. Similarly, the nutritional needs of teenagers change over time, and if teens are using calorie counting or diet-related features, the nutritional goals will need to be periodically adjusted to keep the advice safe and accurate. The device itself should periodically prompt for such updates, perhaps on the teen’s birthday.

In addition, if teenagers are visiting their primary care physician for regular preventative health, the physician could remind or offer to update these body-related settings. But currently, most pediatricians do not have any easy access to the data from the wearable fitness devices of their patients. Ideally, they would have the data and tools to help provide targeted fitness advice (e.g. make sure to walk more on weekends), beyond just adjusting the settings.

Despite the obviously small sample size, it is still possible to draw some early conclusions to potentially inform recommendations. We noted several other issues for wearable device use in the pediatric age range. The teenage female had a contiguous gap in measurements of 7 months. For many years, there was no reminder or warning issued when measurements were not being synchronized from the device. This could easily happen if the synchronization device (i.e. a smartphone or tablet or home computer) is upgraded or restored from a backup. Measurements will be lost if devices are not synchronized periodically. For the Fitbit One used here, the on-device memory only stores 30 days of measurements ^21^. Users should ideally receive a warning if device measurements have not been synchronized to their phones after several days. Fitbit reportedly made an improvement to address this problem in March 2018 ^22^. However, to our knowledge, parents still do not get notifications when their child’s device is not syncing. This should be addressed before long term pediatric use is recommended.

Similarly, teens (or potentially even younger users) will need to remember to periodically charge the device. This habit is harder to adopt, as these devices are worn while sleeping, when other devices (such as phones) are typically charged. Alternative times for consistent charging merit exploration (e.g. while bathing). If the teen has different devices to track steps and sleep, each device will need to be charged when it is not in use.

In this study, we noted the teenager completed fewer steps than the adult. This may be due to a deliberate effort of the adult deliberately to walk at his workplace, whereas the teenager sits in the classroom for most of the day and only walked between classes. However, we noted both individuals walked more during these school/workdays compared to weekend days or holidays. Employers and schools could design layouts that enhance walking activity during the day.

School course schedules are difficult to arrange logistically, but consecutive classes could be separated away from each other on campus, thereby requiring students to walk more between classes. In the workplace, conference calls via mobile devices could be encouraged, thus potentially enabling walking during the calls.

The motivating factors used by the companies making wearable fitness devices can also be tailored for teens. Smartphone-based reminders to exercise could be tailored around teenagers’ unique schedules, with bursts of activity potentially clustered around sports schedules and with periods of less exercise during exam periods.

Anecdotally, the teen started to use the Fitbit One device because she saw her father, the adult in this study, using his device, and because she wanted to be healthier. The prime reason she used the device was to measure her sleep each night, as she could not easily estimate it just by looking at a clock. She could also look at the Fitbit app to see how often she woke up during the night and judge her sleep quality. Instead of relying on qualitative assessments of her lifestyle, she could get quantitative assessments for both her exercise and sleep. As a separate reason, she also used her device because she could see her step count change over time. For example, she could see when she was walking fewer steps per day in middle and high school than she had walked in elementary school. However, the step count was not the primary motivator for continued use of the device. She continued use of the device it because it was easy to integrate into her lifestyle and did not disrupt her daily routine. She developed a habit of charging the device when bathing.

There are several limitations to this study. Obviously, only two individuals were studied here. Others have already been listed above, such as some missing data due to the lack of any device-issued warning. Fewer days of sleep measurements were made, compared to step counts. The Fitbit One needs to be worn while in bed, and requires a button to be manually held down as one is ready to sleep. The same button must then be held down right after waking up to stop the timer. It was common for both individuals to forget to perform either of these two manual steps. A more automated way to determine sleep start and end would be ideal and would enhance accuracy. It is possible that some of these missing measurements might have biased the analysis, if the data were not missing at random.

## Conclusion

Regardless of the limitations, this pilot case study shows that wearable fitness devices can be useful in tracking the long-term health of both adults and teenagers, and we hope to see more studies conducted on the long-term use of these devices.

## Data Availability

The sleep and step count data for the adult and teen used in this analysis are publicly available through Github: https://github.com/kimibutte/Fitbit-Data.

## Acknowledgements

We would like to thank The Harker School for providing the Minitab Express software that was used in analyzing the data. We would also like to thank Chris Spenner (Harker School) for research mentorship and Dr. Tarangini Deshpande (NuMedii, Inc.) for comments on the manuscript.

## Author contributions

Conceptualization, K.D.B., A.J.B., and M.P.S.; Software, A.J.B.; Analysis, K.D.B., A.B., X.L.; Resources, M.P.S.; Writing, K.D.B., A.J.B., X.L., and M.P.S.; Visualization, K.D.B., and A.J.B.; Supervision, X.L., and M.P.S.

